# Monitoring seizure cycles with seizure diaries

**DOI:** 10.1101/2024.08.28.24312758

**Authors:** Ashley Reynolds, Rachel E. Stirling, Samuel Håkansson, Philippa Karoly, Alan Lai, David B. Grayden, Mark J. Cook, Ewan Nurse, Andre Peterson

**Affiliations:** Department of Biomedical Engineering, The University of Melbourne, Melbourne, Australia; Department of Medicine, The University of Melbourne, Melbourne, Australia; Department of Neurosciences, St. Vincent’s Hospital, The University of Melbourne, Melbourne, Australia; Graeme Clark Institute, The University of Melbourne, Melbourne, Australia; Seer Medical, Melbourne VIC 3000, Australia; Department of Clinical Neuroscience, Institute of Neuroscience and Physiology, Sahlgrenska Academy, Gothenburg University, Gothenburg, Sweden

**Author notes:** **Corresponding Author:** Ashley Reynolds, Department of Biomedical Engineering, University of Melbourne, Victoria 3010, Australia. **Co-author(s) details:** Rachel E. Stirling, Samuel Håkansson, Philippa Karoly, Ewan Nurse, Alan Lai, David B. Grayden, Mark J. Cook, Andre Peterson.

**Keywords:** epilepsy, rhythms, periodic, biomarker

## Abstract

**Objective:** The periodicity of seizures, ranging from circadian to circannual cycles, is increasingly recognized as a significant opportunity to advance epilepsy management. Current methods for detecting seizure cycles rely on intrusive techniques or specialised biomarkers, limiting their accessibility. This study evaluates a non-invasive seizure cycle detection method using seizure diaries and compares its accuracy with cycles identified from intracranial electroencephalography (iEEG) seizures and interictal epileptiform discharges (IEDs).

**Approach:** Using data from a previously published first in-human iEEG device trial (n=10), we analysed seizure cycles identified through diary reports, iEEG seizures and IEDs. Cycle similarities across diary reports, iEEG seizures and iEDs were evaluated at periods of 1 to 45 days using spectral coherence, accuracy, precision and recall scores.

**Main results:** Spectral coherence of the raw signals averaged over frequencies and participants indicated moderately similar frequency components between diary seizures/day and iEEG seizures/day (Median=0.43, IQR=0.68). In contrast, there was low coherence between diary seizures/day and IEDs/day (Median=0.11, IQR=0.18) and iEEG seizures/day and IEDs/day (Median=0.12, IQR=0.19). Mean accuracy, precision and recall of iEEG seizure cycles from diary seizure cycles was significantly higher than chance across all participants (Accuracy: Mean=0.95, SD=0.02; Precision: Mean=0.56, SD=0.19; Recall: Mean=0.56, SD=0.19). Accuracy, precision and recall scores between seizures cycles using diary or iEEG compared to IED cycles did not perform above chance, on average. Recall scores were compared across good diary reporters, under-reporters and over-reporters, with recall scores generally performing better in good reporters and under-reporters (Mean=0.74, SD=0.02) compared to over-reporters (Mean=0.37, SD=0.06).

**Significance:** These findings suggest that iEEG seizure cycles can be identified with diary reports, even in both under- and over-reporters. This approach offers a practical, accessible alternative for monitoring seizure cycles compared to more invasive methods.

**Key points:**

1. Seizure cycles identified from seizure diaries can identify the same cycle periods to those detected by intracranial electroencephalography with high accuracy.
2. Seizure under and overreporting reduces the accuracy of seizure cycle detection from diaries.

## 1. Introduction

The periodic occurrence of epileptic seizures has been reported throughout history, with recent advancements in technology able to quantitatively delineate these as seizure cycles^1^. With an estimated 80-90% of people with epilepsy having circadian seizure cycles^2, 3^, 60% multidien^2, 4^, and 12% circannual cycles^4^ there is a major research focus on investigating the utility of cycles for the management of epilepsy. Periodicity assists with forecasting unpredictable seizures, which assists with planning to improve quality of life^5^. There is also potential for these cycles to be used to time short term electroencephalography (EEG) monitoring to improve diagnostic yield^6^, for timing medication^7^, or to support the monitoring of anti-seizure medication efficacy^8^.

Currently, there are several methods that have been used to detect seizure cycles. These methods either use seizure times^6, 9^, or EEG waveforms that are specific epilepsy biomarkers such as interictal epileptiform activity^4, 8, 10–16^ and high frequency oscillations^17^. However, such biomarkers are not present in everyone with epilepsy^18–21^ and are not always a good marker for seizure timing^2, 11, 22–24^. Furthermore, continuous monitoring of epilepsy biomarkers presently requires invasive implanted devices like intracranial or sub-scalp EEG, though these are not yet widely available due to regulations and accessibility. Other potential physiological biomarkers of seizure cycles, such as resting heart rate, have been identified^25, 26^, but are not well understood in the context of epilepsy and seizure risk.

Due to the sparsity of seizures over time, cycle detection methods using seizure times require long durations of data to estimate the cycle period, which then informs fixed-period sinusoids to model the phase of seizure risk. However, seizure cycles appear almost-periodic, fluctuating between a range of periods^27^. Thus, using all available data and a fixed-period means previous methods that rely on seizure times do not perform as well as irregular cycles of interictal epileptiform activity when forecasting seizures^12^. However, cycles of interictal epileptiform activity are not a perfect seizure cycle biomarker and methods based on them suffer from various patient-specific rates of false positives and false negatives^4, 11, 13, 15, 27^. This highlights the problem that there is no perfect method to detect seizure cycles.

Improving upon a non-invasive diary-based cycle detection method would be highly desirable, as it could widen the clinical application of seizure cycles. Particularly because seizure diaries are a cornerstone of clinical practice and heavily relied upon for monitoring epilepsy and treatment response^28^. However, the accuracy of seizure diaries is highly variable between individuals, depending on seizure type, the state from which they occur (awake, drowsy, or asleep), the type of diary (electronic or paper) and who is making the report (patient or caregiver)^29–35^. Studies using video EEG to verify diary accuracy indicate 23-38% of people underreport seizures and of these people, 27-100% of seizures are not reported^29–31, 33–35 36–40^. Similar studies have also demonstrated 38-57% of patients overreport seizures, with 58% of all reported events possibly being non-seizure events of uncertain origin^30, 35^. The inaccuracy of self-reported seizures causes inaccurate estimates of circadian seizure cycles^32^. However, evidence exists that multidien seizure cycles can be identified from diaries, despite their inaccuracies^6, 41–43^.

Therefore, our aim is to investigate an improved seizure cycle detection method based on seizures diaries and compare its ability to identify similar cycles to those identified by both intracranial EEG (iEEG) detected seizures and interictal epileptiform discharges (IEDs). We also aim to compare the similarities between methods when participants are stratified as good reporters, under-reporters and over-reporters. We hypothesise that people who self-report seizures with reasonable accuracy will have similar seizure cycles detected by both diary and by iEEG. However, the detected cycles will be less similar in people who under- or over-report seizures. We also hypothesise cycles detected using iEEG seizures will identify similar cycle periods to IED cycles, but they will not be identical due to the variable relationship between interictal epileptiform discharges and seizures^22, 23^.

## 2. Methods

### 2.1 Study design

This study used the previously published NeuroVista dataset, a first-in-human longitudinal clinical trial of an implantable seizure advisory device for people with epilepsy^33^. Using this dataset, we compared three methods to identify seizure cycle periods: 1. diary seizure cycles, defined as fixed-period cycles applied to sliding windows of participants’ self-reported seizure diaries, 2. iEEG seizure cycles, defined as fixed-period cycles applied to sliding windows of participants’ iEEG seizure records, and 3. IED cycles, identified using a wavelet transform applied to IEDs per hour. Strength of candidate cycles were defined by the synchronisation index (SI) values of seizure times mapped to phases of the cycle.

### 2.2 Participants

Trial participants were from Melbourne, Australia. They had drug-resistant focal epilepsy and were consistently taking the same anti-seizure medications and doses throughout the original trial. Participants were included in this study if they had >10 self-reported and iEEG seizures and >6 months of recording between 2010-2012 (n=10 of 15).

### 2.3 Seizure and interictal epileptiform discharge detection

Seizures and IEDs were confirmed by board-certified epileptologists. Both clinical and sub-clinical seizures (iEEG seizures associated with and without clinical symptoms, respectively) were included. We refer the reader to the original study^33^ for details.

### 2.4 Pre-processing iEEG data

Hourly counts of IEDs were used to detect IED cycles. For every hour of the total iEEG recording, the proportion of missing data during that hour was recorded. If the proportion of missing data was less than or equal to 50%, then the number of IEDs was normalised by this proportion^2^. Any hour where the proportion of missing data was greater than 50% was considered too unreliable and treated as completely missing. If there were adjacent hours of completely missing data, where the total duration was less than 20% of the cycle period of interest, then hourly IED values were imputed^12^. This involved linear interpolation between the means of the adjacent segments of data. The length of the adjacent segments was either five times the length of the missing segment or available data up to the nearest missing segment (whichever was shorter)^12^. For example, a missing segment of 2 hours was estimated from the means of adjacent segments up to 10 hours either side of the missing segment. Gaussian white noise with a standard deviation from the concatenated adjacent segments was added to the imputed values^12^. Hours of completely missing data lasting longer than 20% of the cycle period of interest were not interpolated and instead data before and after the gap were analysed independently^15^.

### 2.5 Cycle comparison

#### 2.5.1 Spectral coherence of signals

We first investigated whether diary seizures/day, iEEG seizures/day, and IEDs/day share frequency components using spectral coherence. The magnitude-squared coherence provides a value between 0 and 1 for each frequency (i.e., inverse of period), indicating a level of correlation. A value of 0 signifies no correlation between the signals at a given frequency, while a value of 1 indicates perfect correlation. Statistically significant coherence values for each frequency were determined using a permutation test. In this test, each time-series was randomly shuffled to create 1000 permutations. We computed spectral coherence values for frequencies corresponding to cycle periods ranging from 0 to 45 days^15^. A 5% false discovery rate was set as the threshold for determining the critical coherence value.

#### 2.5.2 iEEG and diary seizure cycle detection

Seizure cycle detection using seizures employs phase-locking to identify cyclical patterns in noisy data^2^. Phase-locking is the tendency for events to occur during a small range of phases in a cycle and can be measured by the synchronisation index (SI) value for a given cycle period. To calculate SI values, seizure start times were represented on a unit circle (Figure 1). The circular average of the seizure times is the mean phase vector (MPV), with a magnitude between 0-1 and phase from 0 to 2π. The magnitude gives the SI value. A magnitude of 1 indicates seizures are perfectly phase-locked to a cycle with *x* period indicating seizure likelihood is certainly modulated by a cycle with *x* period^2^. A magnitude of 0 indicates seizures occur uniformly between 0 to 2π and is improbable that a cycle with *x* period modulates seizure timing. The phase of the MPV indicates where in the cycle seizures are more likely to occur.

**Figure 1.**
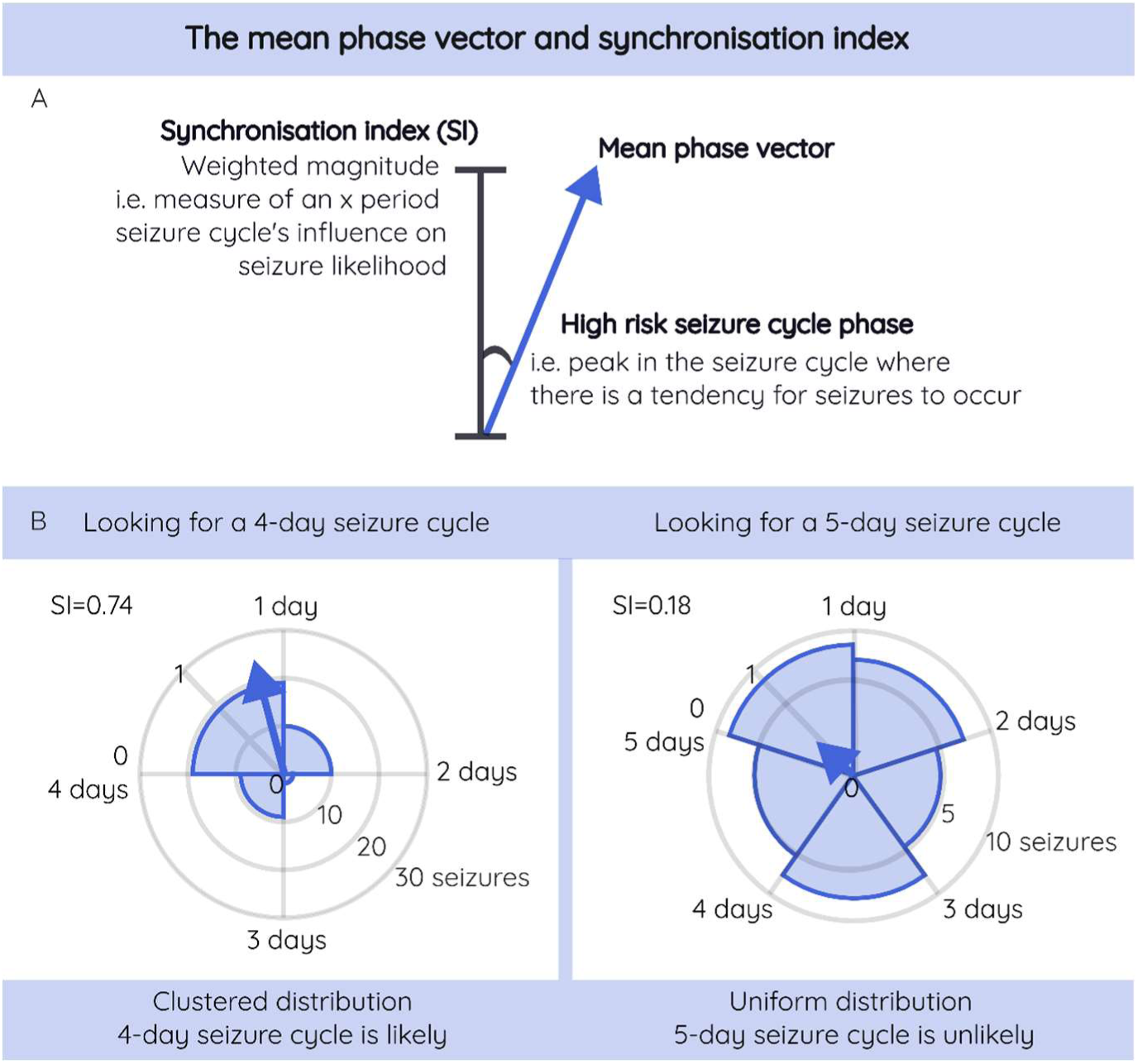
The mean phase vector (MPV) and synchronisation index (SI). (A) The weighted magnitude of the MPV, along the radial-axis of the unit circle, represents the SI value and indicates how influential this cycle period is in modulating seizure likelihood (effect size). The phase of the MPV indicates where in the cycle seizures have a tendency to occur. (B) An example of detecting a 4-day seizure cycle using the SI method. On the left side, seizure times are grouped into 4 1-day intervals on a circular histogram (note, binning is only used for visualisation purposes and not used in the actual method). The average of the histogram is the arrow, the MPV. Seizures occur clustered around day 1 of the 4-day cycle and the SI is close to 1, indicating seizure likelihood is probably modulated by a 4-day cycle. On the right side, seizure times are grouped into 5 1-day intervals. Seizures occur uniformly around the circle and the SI is close to 0, indicating seizure likehood is probably not modulated by a 5-day cycle. It should be noted that seizure rate will negatively influence the SI, as seizure rate affects the density of seizures occurring around the unit circle. In this example, one seizure every 4 days produces less densly distributed seizures than one seizure every 5 days. The renormalisation factor adjusts for this bias giving the SI value^45^.

Unlike previous methods^2, 6, 9^, all seizures, instead of only the first seizure in a cluster, were included to calculate SI values in this work. This modification was justified as there is no single definition of a seizure cluster^44^. A renormalisation factor described by Andrzejak et al., (2023) was then applied to account for the number of seizures influencing the SI^45^ and changing the range of SI values to be ≤0-1. Although this modification helps to reduce the number of seizures biasing the SI, it does not completely account for cycle period bias. That is, shorter cycles are sampled more frequently than longer cycles. More repetitions of a cycle are akin to repeating experiments, which can smooth out fluctuations due to random noise and outliers. As the time for any analysis is finite, longer seizure cycles will always be under sampled compared to shorter cycles.

For each cycle, the SI was defined as:

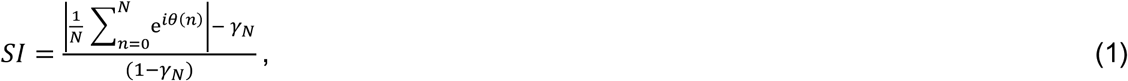

where *N* is the total number of seizures, *n* is the seizure number in sequence, *θ* is the phase of the *n*^th^ seizure relative to the cycle of interest, and *i* is the imaginary number. *γ*_N_ is a renormalisation factor^45^:

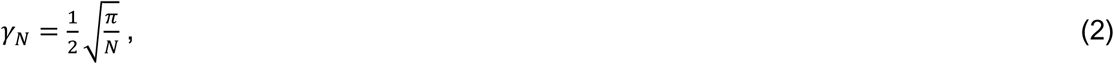

SI values of seizure cycles were evaluated at periods between 1 and 45 days. To identify whether a circadian cycle exists, only a 24-hour period was considered. For multidien cycles, only integer cycle periods between 2 days to 45 days were considered^15^, using integer values with 1-day intervals. Integers were used to reduce computations and simplify the method for clinical use. Although this introduces inaccuracy for cycle periods fluctuating between integer days, 1-day intervals are more practical for clinical implementation where clinical review periods occur over months. If greater precision is needed, cycles that are fractions of a day could be included in future studies with sufficient computational resources.

Of the 45 possible seizure cycles periods, only those that were statistically likely to influence seizure probability were considered. This was based on a permutation test that set a period-specific SI threshold, to mitigate the period bias in the SI^46, 47^. Briefly, 1000 permutations of the null hypothesis, one without periodicity, were generated for each window of data by randomly shuffling seizure start times with their associated inter-seizure interval^46, 47^. SI values were calculated and a false discovery rate of 10% was used as the SI threshold to identify statistically significant seizure cycle periods^4^. Statistically significant periods that were within ±33.33% of a central period with peak SI, were assumed to be the same cycle and so only the period with the peak SI was considered^15^.

An approximate 135-day sliding window of data was used to detect seizure cycles over time. The window length was selected due to the sparsity of seizure data and to allow cycle periods to repeat a minimum of 3 times within an approximate 4-month clinical review period^28^. We slid the window by the number of days equivalent to the cycle period of interest to prevent an incomplete cycle inaccurately influencing the MPV and reran the analysis iteratively^15^ (Figure 2).

**Figure 2.**
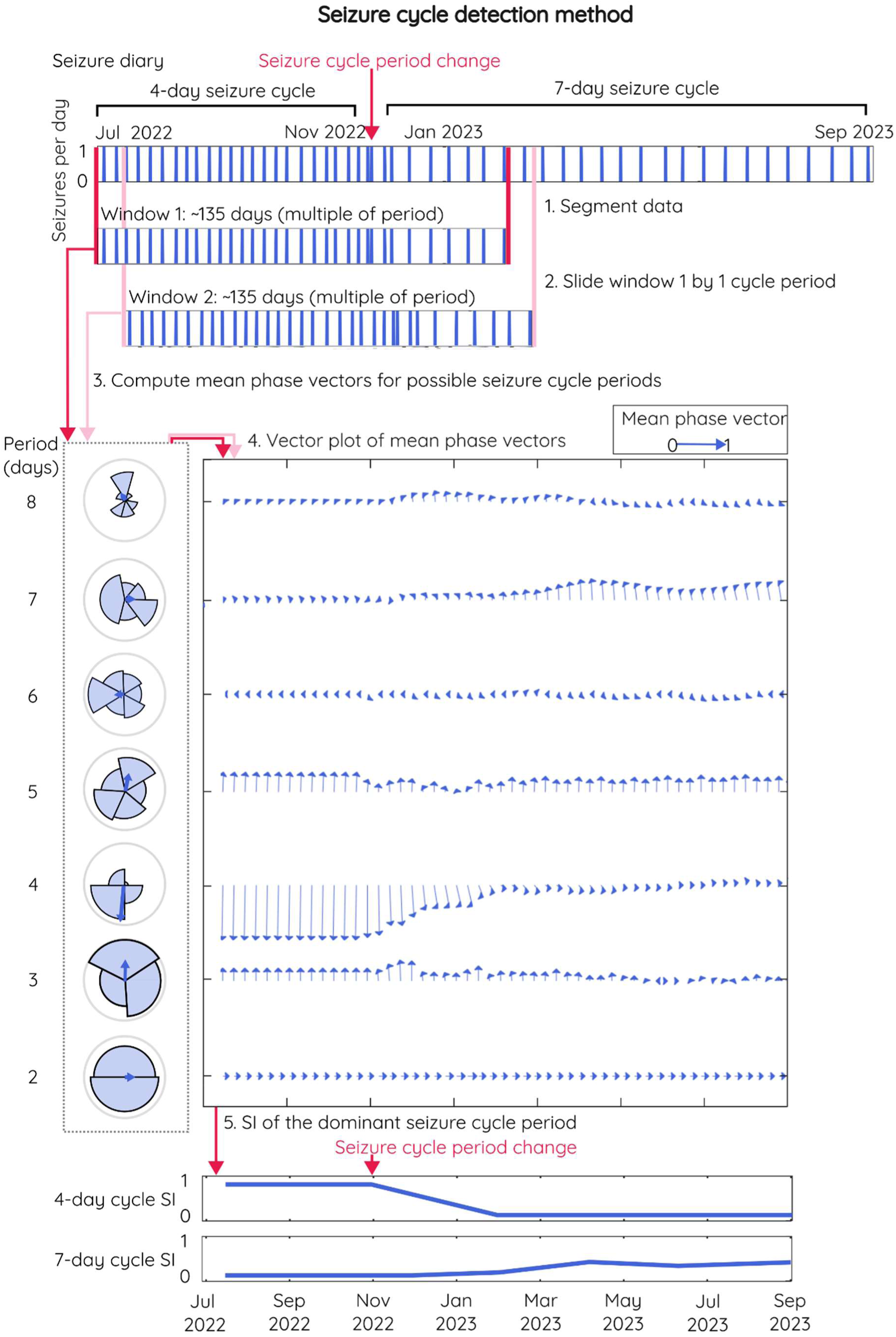
Seizure cycle detection method. To demonstrate the method, a simple simulated seizure diary is used. The diary has seizures initially occurring on average every 4 days. Three months later it changes to every 7 days. The first approximate 135-days of the diary are divided into the possible cycle periods of interest (in this example, 2–8-day cycles) and plotted as circular histograms (for visualisation only) and the weighted circular average is the mean phase vector (MPV), represented as an arrow. The MPVs for all example cycle periods are plotted in the first column of the vector plot (y-axis). The window of data moves by one period length of interest and the analysis is re-run. The recalculated MPVs are plotted in the next column(s). E.g., for a 5-day cycle, the window moves by 5 days, and the 5-day cycle MPV is calculated and the same MPV is plotted repeatedly for the next 5 days along the x-axis. With each iteration, the change in the average phase (angle of the arrow) and magnitude (length of the arrow, synchronisation index (SI)) can be tracked over time (x-axis). However, for visualisation purposes, arrows were down sampled and plotted every 7 days. Note, tiny vectors (seen for example at 6-days) indicate the SI values are close to 0, suggesting a very low likelihood that a cycle exists at this period. In the first 3 months the vectors are greatest at the 4-day cycle, indicating a 4-day cycle is likely. In the following months, the 4-day cycle vectors reduce in magnitude and the phase changes, the 7-day cycle vectors increase in magnitude and the phase changes; together this indicates a change in the dominant seizure cycle period and effect size. It is also important to observe the magnitude of the vectors of the 4-day cycle are always greater than the sub-harmonic and harmonic cycle periods (2 and 8 days, respectively, in this example) in the first 3 months.

#### 2.5.3 Interictal epileptiform discharge cycle detection

IED cycles were identified using continuously recorded iEEG-detected IEDs. A continuous analytic Morse wavelet transform of IEDs per hour was implemented^15^. The maximum period investigated was defined by the length of the data, but only integer periods of 1-45-days were included for methodological comparisons. A permutation test to identify significant cycles was used by randomly shuffling hourly IED values 1000 times. A threshold of the magnitude of the wavelet coefficient was calculated to identify significant cycles using a false discovery rate of 10%. Significant cycles were defined as peaks in the wavelet periodogram whose power values were above the significance threshold. Like seizure cycle detection, for two or more significant cycles within proximity (within a ±33.33% range) of each other, only the period with the peak power value above the threshold was accepted as the significant cycle.

### 2.6 Comparing cycles detected by each method

Pairwise comparisons between diary seizure cycles, iEEG seizure cycles and IED cycles were made using accuracy (the number of true positives and true negatives out of all the total sample), precision (the number of true positives out of the number of true positives and false positives) and recall (the number of true positive out of the number of true positives and false negatives) scores. iEEG seizure cycles were used as the ground truth to compare diary seizure cycles and IED cycles. When comparing diary cycles to IED cycles, IED cycles were used as the ground truth.

Accuracy, precision and recall values of the cycle array (positive/negative cycle detection for each possible period) were calculated each day of the recording. The mean accuracy, precision and recall values across all days were reported for each participant. Statistical significance of accuracy, precision and recall values was determined using a permutation test. For each participant, 1000 permutations were generated each day by randomly shuffling the daily cycle array (i.e., keeping number of positives and negatives consistent) and the 90^th^ percentile was used as the significance threshold (i.e., 10% false discovery rate). Mean significance thresholds across all days of the recording were calculated, and accuracy, precision and recall values above this threshold were determined to be significantly higher than chance.

It should be noted that the IED cycle method is a continuously sampled time-series with IED counts every hour. This allows for very flexible window sizes to calculate each scale magnitude for the wavelet transform, providing greater temporal resolution to detect cycle periods. Comparatively, this seizure cycle method does not have as fine-grained temporal resolution, as it maintains a relatively large window for sparse seizure data (more zero-seizure days than seizure days). Due to the different windowing methods, it is not expected that the identified cycles align at each timepoint, but we expect that we should observe alignment some of the time.

For a straightforward interpretation of results from spectral coherence and the performance metrics (accuracy, precision and recall), we categorised the values into five bins: very low (0.00-0.20), low (0.21-0.40), moderate (0.41-0.60), high (0.61-0.80), and very high (0.81-1.00).

### 2.7 Effect of overreporting and underreporting on cycle detection

To assess the effect of overreporting and underreporting on seizure diary cycle detection, participants were stratified by the accuracy of their self-reported seizures and the performance metric, recall was reported. All events, which included self-reported seizures with iEEG (i.e., electrographic) seizure correlate (correctly reported), iEEG seizures without self-reported seizure correlate (underreported), and self-reported seizures without iEEG correlate (overreported) were totalled. Participants were ‘good reporters’ if >95% of the total events were correctly reported. ‘Rare’ under or overreporting was defined as ≤5% of the total number of events were seizures without self-reported seizure correlate or self-reported seizures without iEEG correlate, respectively. ‘Occasional’ under or overreporting was defined as >5-≤10% of events were under or overreported. ‘Sometimes’ was >10-≤25% of events were under of overreported. Often was defined as >25-≤50% of all events were under of overreported. ‘Frequent’ was >50% of all events were under or overreported.

All analyses were undertaken using MATLAB (v2024b, MathWorks Inc., Natick, MA, USA) and the Circular Statistics Toolbox^48^.

## 3. Results

### 3.1 Spectral coherence

Figure 3 shows spectral coherence values per candidate period across comparison pairs of IEDs/day, iEEG seizures/day and diary seizures/day. When comparing IEDs/day and iEEG seizures/day, 9 out of 10 participants (all but S3) showed a small proportion of statistically significant correlations in the frequency domain, although not all periods were highly correlated. By observation, periodic components of IEDs/day and iEEG seizures/day of 20 days or less tended to be more similar than longer periods. Median spectral coherence values between IEDs/day and iEEG seizures/day were 0.12 (IQR=0.19) across all participants and candidate periods. A similar trend in spectral coherence was observed when comparing IEDs/day and diary seizures/day (Median=0.11, IQR=0.18). However, any differences between the two spectral coherence trends for IEDs/day and seizures/day were due to low correlations between iEEG seizures/day and diary seizures/day at respective periods. Overall, the spectral density of diary and iEEG seizures were similar, with moderate to high spectral coherence values (Median=0.43, IQR=0.68). Notably, 4 out of 10 participants had coherence values between 0.90 and 1.00 for periods longer than 20 days, suggesting that the periodic components of both seizure recording methods were almost identical in these participants (see Figure 3).

**Figure 3.**
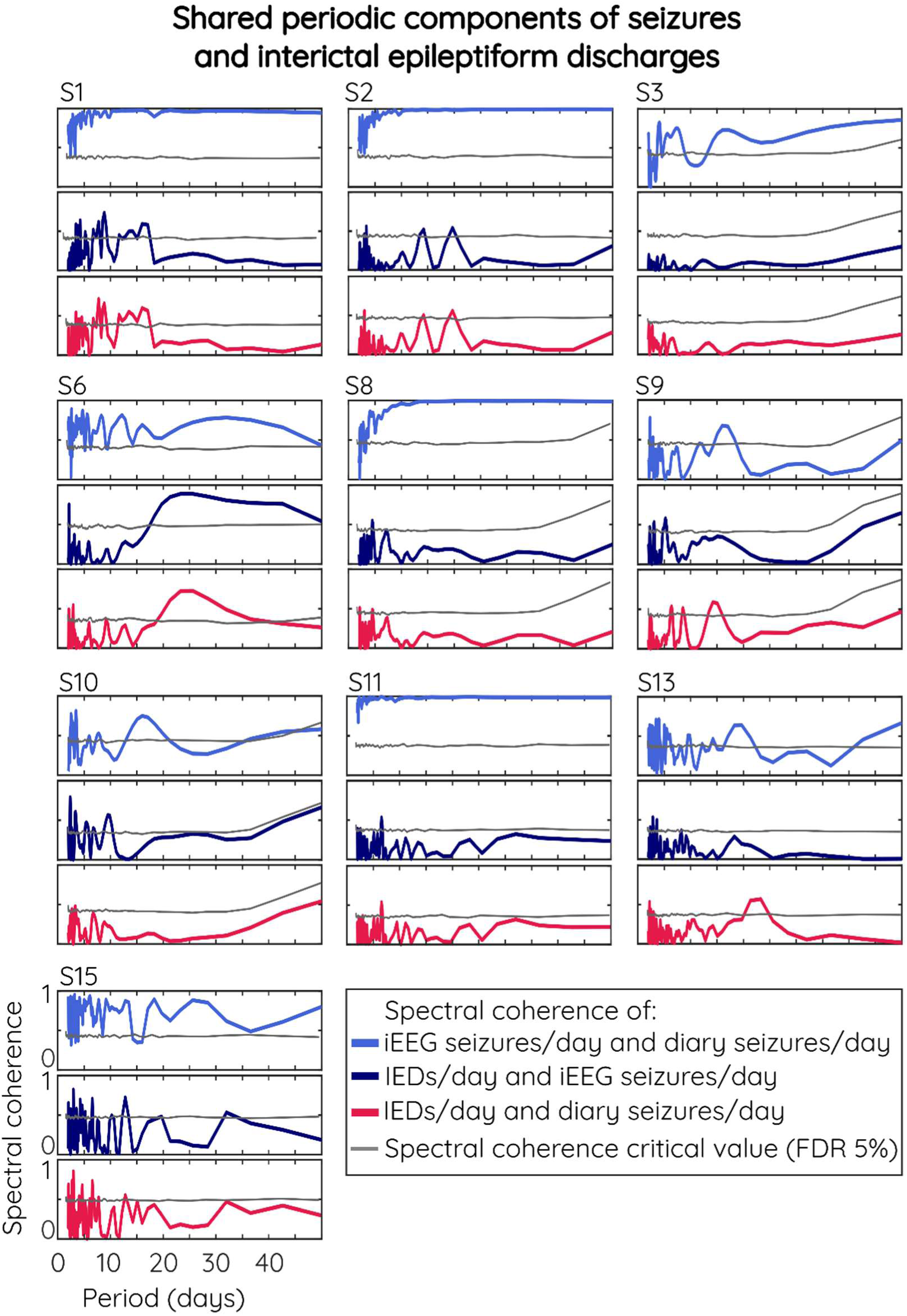
Periodic components are similar but not identical between diary seizures/day iEEG seizures/day and IEDs/day. Periodic components of IEDs/day were less similar to both iEEG and diary seizures/day. Similarities were also patient specific but tended to have more similar frequency components for shorter periods. Spectral coherence values above the critical value indicate statistically significant correlations. FDR: false discover rate.

### 3.2 Comparing cycle detection methods

The seizure diary method produced similar cycle results to the iEEG seizure cycle method over 1-2 years (Table 1). When comparing iEEG seizure cycles to diary seizure cycles, the mean accuracy across all participants was 0.95 (SD=0.02), whereas both precision and recall scores had a mean of 0.56 (SD=0.19) across all participants. The moderate to high accuracy values were a reasonable result given the spectral coherence measures demonstrated the periodicity between the different signals were not perfectly aligned and were also patient specific.

**Table 1.** Seizure cycles detected using seizure diaries identify similar cycles to those detected by iEEG but are less similar to IED cycles. The performance of diary seizure cycles and IED cycles compared to iEEG seizure cycles, assessed by accuracy, precision and recall. ^a^ground truth. *significantly better performance than 90% of random permutations.

|  | Accuracy |  |  | Precision |  |  | Recall |  |  |
| --- | --- | --- | --- | --- | --- | --- | --- | --- | --- |
| Participant | iEEG<sup>a</sup> and diary | iEEG<sup>a</sup> and IEDs | IEDs<sup>a</sup> and diary | iEEG<sup>a</sup> and diary | iEEG<sup>a</sup> and IEDs | IEDs<sup>a</sup> and diary | iEEG<sup>a</sup> and diary | iEEG<sup>a</sup> and IEDs | IEDs<sup>a</sup> and diary |
| 1 | 0.96* | 0.91 | 0.91 | 0.70* | 0.23 | 0.19 | 0.70* | 0.20 | 0.22 |
| 2 | 0.97* | 0.91 | 0.91 | 0.76* | 0.25 | 0.23 | 0.78* | 0.23 | 0.25 |
| 3 | 0.94* | 0.91 | 0.92* | 0.52* | 0.26 | 0.25 | 0.44* | 0.25 | 0.21 |
| 6 | 0.93* | 0.91 | 0.91 | 0.42* | 0.16 | 0.11 | 0.44* | 0.13 | 0.15 |
| 8 | 0.97* | 0.90 | 0.90 | 0.75* | 0.10 | 0.05 | 0.74* | 0.09 | 0.05 |
| 9 | 0.92* | 0.91 | 0.90 | 0.32* | 0.10 | 0.06 | 0.35* | 0.07 | 0.09 |
| 10 | 0.92* | 0.91 | 0.91 | 0.30* | 0.18 | 0.24 | 0.33* | 0.19 | 0.23 |
| 11 | 0.97* | 0.91 | 0.91 | 0.75* | 0.17 | 0.16 | 0.74* | 0.17 | 0.15 |
| 13 | 0.92* | 0.90 | 0.90 | 0.37* | 0.14 | 0.13 | 0.36* | 0.14 | 0.14 |
| 15 | 0.96* | 0.90 | 0.90 | 0.71* | 0.07 | 0.06 | 0.70* | 0.06 | 0.06 |
| Average ± | 0.95* ± | 0.91 ± | 0.91 ± | 0.56* ± | 0.17 ± | 0.15 ± | 0.56* ± | 0.15 ± | 0.16 ± |
| SD | 0.02 | 0.01 | 0.01 | 0.19 | 0.07 | 0.08 | 0.19 | 0.07 | 0.07 |

Comparing the cycle periods detected by seizure diary and IEDs, the accuracy was 0.91 (SD=0.01) with poor precision (Mean=0.15, SD=0.08) and recall (Mean=0.16, SD=0.07). However, this was similar when comparing iEEG seizures cycles and IED cycles (accuracy Mean=0.91, SD=0.01, precision Mean=0.17, SD=0.07, and recall Mean=0.15, SD=0.07). These low values can be expected when put into context with the spectral coherence results, which demonstrated limited similarity between the periodicity of seizures (both iEEG and diary) and IEDs.

### 3.3 Effect of overreporting and underreporting on cycle detection

The single good reporter who rarely underreported seizures had a recall of 0.78 for iEEG seizure cycles compared to diary seizure cycles (Table 2). Their recall value for iEEG and diary seizure cycles compared to IED cycles was also slightly higher compared to the rest of the cohort. Categorising participants by under and overreporting, indicates that frequent overreporting reduces recall values between the diary seizure cycles and iEEG seizure cycles. Underreporting of any frequency, rare through to frequent (with only rare or occasional overreporting) produced average recall values between 0.70-0.78 of the diary seizure cycles compared to iEEG seizure cycles.

**Table 2.**
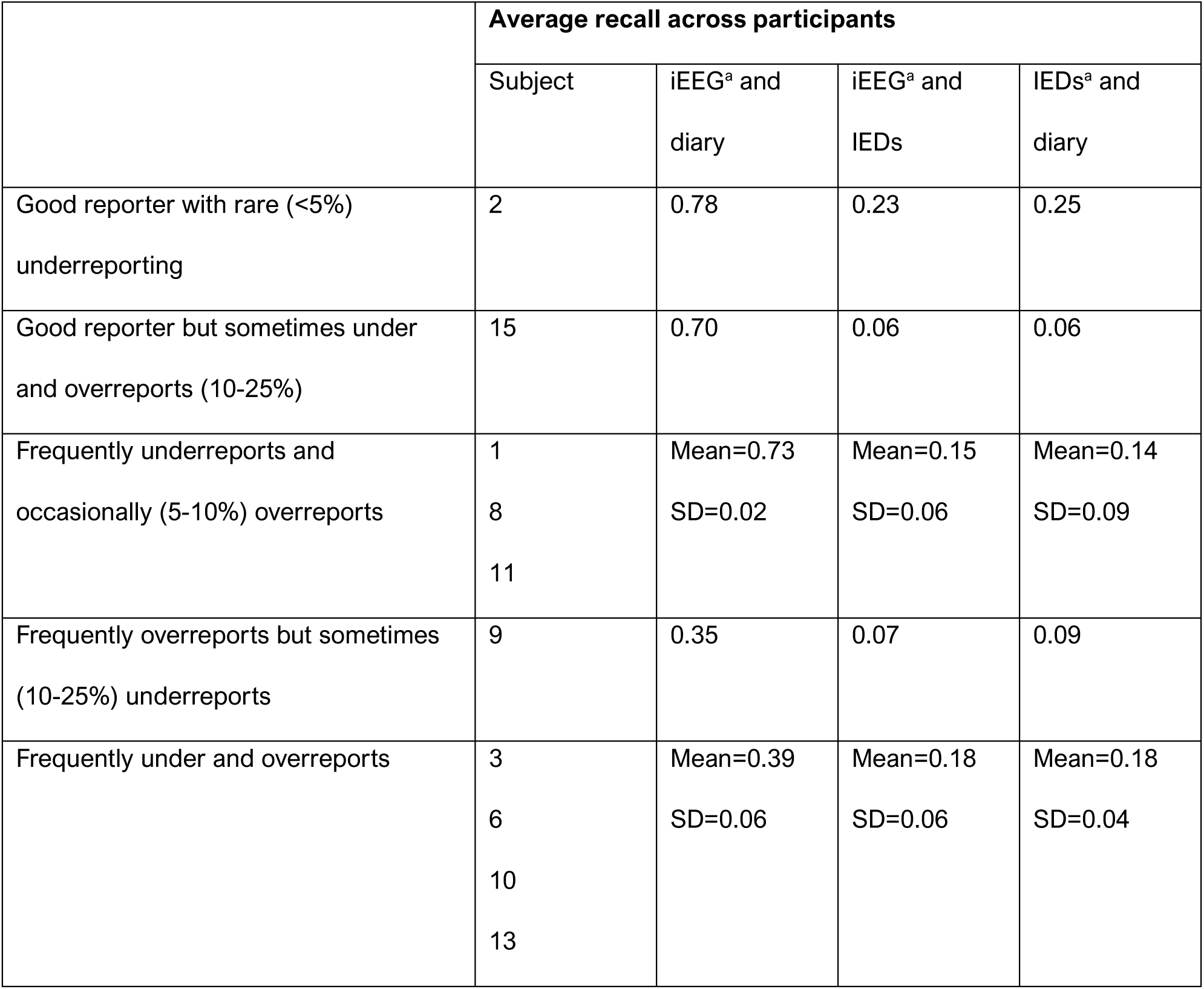
Frequent under or overreporting negatively affects seizure cycle detection. Performance as assessed by recall, of methods to detect seizure cycles stratified by participants who under and/or overreport seizures.

## 4. Discussion

This study demonstrates that a non-invasive diary-based method can detect seizure cycles of 1-45 day period lengths with similar accuracy compared to iEEG detected seizure cycles. As hypothesised, the level of accuracy was patient specific and depends on the amount of under and overreported seizures. Moreover, when comparing the seizure cycles that were detected by diary or iEEG, both detection methods intermittently identified similar cycle periods compared to the IED method. This was limited and may be attributed to the general low coherence observed between the periodicity of the original seizures and IED signals.

An accessible seizure cycle detection method is crucial for maximizing the potential benefits offered by seizure cycles. Although IED cycles can identify cycles of seizure risk, IED detection is unfortunately restricted to a small subset of people who have IEDs, as they are not present in every person with epilepsy^18–21^. There is also significant intra- and inter-individual variability in the relationship between IEDs and seizures^2, 11, 22, 23^, which our study agreed with.

It is possible that the discrepancies between IEDs and seizure cycles^11^ lie in the limitations of iEEG devices. There are a small number of studies investigating the relationship between IEDs and seizure timing over long timescales in participants using the NeuroVista device^22, 49^ and sub-scalp EEG systems^11, 16, 41^. However, most of the previous research in this field was conducted with the Responsive Neurostimulation (RNS) system (NeuroPace, Mountaint View, CA)^4, 12, 13, 15^. Interictal epileptiform activity detected using the RNS system included IEDs plus other waveforms^50^. Further, due to device storage capacity, the device only records interictal epileptiform activity during pre-set times or when triggered^50^. This may result in over-sampling of one cycle phase and under-sampling of others. In addition, RNS system study participants also receive neurostimulation that previous research indicates can alter both IED and seizure cycles^10, 51^. Thus, it remains unknown how a non-specific algorithm with non-uniform sampling and neurostimulation affects seizure cycle detection and generalisability. However, these technological limitations are not the same for all devices and the IEDs detected in this study were specific and uniformly sampled. Therefore, it is likely that IEDs serve as biomarkers with constrained predictive capabilities, rather than representing the true underlying seizure cycle; particularly if there are other biological or environmental cycles influencing the temporal patterns of seizures^1^. This is supported by previous research demonstrating the highly varied, patient-specific relationship between seizure timing and IED counts^22^. Though we found it useful to compare the similarity in cycle results between the IED method and seizure method, it may not be necessary or even appropriate to compare one method to the other.

For people who cannot access the RNS system or other ultra-long EEG system in the future (e.g., because they do not have drug-refractory epilepsy; do not live in the correct location or have financial means; do not meet other specific device, surgical or anaesthetic criteria; or are unwilling to accept the risks of an implant) it is useful to know that diary-based cycle detection can be accurate. Also, it can be used in anyone who can maintain any kind of seizure diary (e.g., paper-based or electronic^35^) with reasonable accuracy (incorrect reports up to 25% of the events). Going forward, the current seizure cycle method could be further optimised by using more flexible window sizes and fractions of a day^6^, to improve the temporal resolution of cycle detection. Diary-cycle detection could also be supplemented by other biological cycle detection that utilise wearable devices to continuously record biosignals such as resting heart rate, heart rate variability, accelerometery, electrodermal activity and peripheral body temperature^25, 52^. It has been demonstrated in a small group of people with focal epilepsy and the RNS system that multiscale cycles present in these biosignals phase-lock with seizures^52^. However, given the small sample size and individual variability between biosignals and seizures, further investigations are required to examine the generalisability of results, as well as delineate any underlying mechanisms driving and linking their temporal structures.

Given we do not know the mechanisms underlying seizure cycles or the gold-standard method to track them, optimising a cycle detection method may be challenging. Comparing an imperfect seizure cycle method to an imperfect IED method can only give an indication of the similarities in detectable cycles. Though realistic seizure time simulators exist^53^, the cyclical component is modelled by non-physiological fixed-period sinusoids. Both options can only provide limited insight into how useful a diary-based cycle detector could be. Until we discover the sources of seizure cycles and how to monitor them, inaccurate cycle assumptions applied to methodological decision will likely limit their clinical utility.

It should be noted that this study was limited by missing data, but interpolation methods were kept to a minimum, consistent with previously published work^4, 10, 15^. Moreover, although we investigated the effects of under and overreporting on cycle period detection, we did not investigate the effects of missing data on phase estimation, which has previously been demonstrated^32, 54^. This study did not investigate phase estimations due to the coarse time window of the seizure cycle method, which was only able to provide an average phase over the approximate 4-month window, thus unlikely to be informative for daily life. Addressing this limitation will require future methodological development.

## 5. Conclusion

A non-invasive diary-based seizure cycle detection method can identify cycles comparable to those found through invasive iEEG recorded seizures. This approach expands opportunities for monitoring seizure cycles, with potential benefits in clinical settings such as refining treatment evaluations, guiding chronotherapy, and providing patients with insights into seizure risk to better understand and manage their condition. Its significant accessibility could prove to be extremely beneficial for individuals with epilepsy who already maintain seizure diaries.

## Acknowledgments

A.R. receives funding from the Australian Government Research Training Program Scholarship from the University of Melbourne.

## Author contributions

A.R. undertook the literature review, designed the methodology, undertook the analysis and results interpretation, wrote the manuscript, and created the figures and tables. R.E.S. assisted with methodological design and analysis, results interpretation, wrote and edited the manuscript. S.H. edited the manuscript. P.K. provided pre-processed NeuroVista data, assisted with interpretation of results and edited the manuscript. E.N. edited the manuscript. A.P., A.L., and D.B.G assisted with methodological design, assisted with interpretation of results, contributed to concepts, edited the manuscript, and supervised A.R. M.J.C. contributed to concepts, provided access to the data, assisted with interpretation of results, edited the manuscript, and supervised A.R.

All authors approved the final version of the manuscript. A.R., P.K., A.L., and M.J.C. had full access to the data. No author was precluded from accessing data in the study.

## Data availability

Deidentified NeuroVista data are available upon reasonable request to co-author Prof. Mark J. Cook.

